# A Randomised -Controlled Phase 2 trial of Molnupiravir in Unvaccinated and Vaccinated Individuals with Early SARS-CoV-2

**DOI:** 10.1101/2022.07.20.22277797

**Authors:** Saye H Khoo, Richard FitzGerald, Geoffrey Saunders, Calley Middleton, Shazaad Ahmad, Christopher J Edwards, Dennis Hadjiyiannakis, Lauren Walker, Rebecca Lyon, Victoria Shaw, Pavel Mozgunov, Jimstan Periselneris, Christie Woods, Katie Bullock, Colin Hale, Helen Reynolds, Nichola Downs, Sean Ewings, Amanda Buadi, David Cameron, Thomas Edwards, Emma Knox, I’ah Donovan-Banfield, William Greenhalf, Justin Chiong, Lara Lavelle-Langham, Michael Jacobs, Wendy Painter, Wayne Holman, David G Lalloo, Michelle Tetlow, Julian A Hiscox, Thomas Jaki, Thomas Fletcher, Gareth Griffiths, the AGILE CST-2 Study Group

**Affiliations:** Pharmacology and Therapeutics, University of Liverpool, Liverpool, UK; Tropical and Infectious Disease Unit, Liverpool University Hospital NHS Foundation Trust, Liverpool, UK; NIHR Royal Liverpool and Broadgreen Clinical Research, Liverpool University Hospital NHS Foundation Trust, Liverpool, UK; Southampton Clinical Trials Unit, University of Southampton, Southampton, UK; NIHR Manchester Clinical Research Facility, Manchester, University NHS Foundation Trust, Manchester, UK; NIHR Southampton Clinical Research Facility, University Hospital Southampton NHS Foundation Trust, Southampton, UK; Human Development and Health School, University of Southampton, Southampton, UK; NIHR Lancashire Clinical Research Facility, Lancashire Teaching Hospitals NHS Foundation Trust; Clinical Directorate, University of Liverpool, Liverpool, UK; MRC Biostatistics Unit, University of Cambridge, Cambridge, UK; NIHR Kings Clinical Research Facility, King’s College Hospital NHS Foundation Trust, London, UK; Molecular & Clinical Cancer Medicine, University of Liverpool, Liverpool, UK; Centre for Drugs and Diagnostics, Liverpool School of Tropical Medicine, Liverpool, UK; Institute of Infection, Veterinary and Ecological Sciences, University of Liverpool, Liverpool, UK; National Institute of Health Research, Health Protection Research Unit in Emerging and Zoonotic Infections, University of Liverpool, Liverpool, UK; Infectious Diseases, Royal Free London NHS Foundation Trust, London, UK; Ridgeback Biotherapeutics, Miami, Florida, USA; Director, Liverpool School of Tropical Medicine, Liverpool, UK; Computational Statistics, University of Regensburg, Regensburg, Germany; Clinical Sciences, Liverpool School of Tropical Medicine, Liverpool, UK

## Abstract

**Background:** Molnupiravir was licensed for treating high-risk patients with COVID-19 based on data from unvaccinated adults. AGILE CST-2 (NCT04746183) Phase II reports safety and virological efficacy of molnupiravir in vaccinated and unvaccinated individuals.

**Methods:** Adult out-patients with PCR-confirmed SARS-CoV-2 infection within five days of symptom onset were randomly assigned 1:1 to receive molnupiravir (800mg twice daily for five days) or placebo. The primary outcome was time to swab PCR-negativity, compared using a Bayesian model for estimating the probability of a superior virological response (Hazard Ratio>1) for molnupiravir over placebo. Secondary outcomes included change in viral titre at day 5, safety and tolerability, clinical progression and patient reported outcome measures. We analysed outcomes after the last participant reached day 29.

**Findings:** Of 180 participants randomised (90 molnupiravir, 90 placebo), 50% were vaccinated. Infections with SARS-CoV-2 variants Delta (40%), Alpha (21%), Omicron (21%) and EU1 (16%) were represented. The median time to negative-PCR was 8 versus 11 days for molnupiravir and placebo (HR=1·30, 95% CrI 0·92-1·71, p=0·07 by Logrank and p=0·03 by Breslow-Gehan tests). Although small numbers precluded subgroup analysis, no obvious differences were observed between vaccinated and unvaccinated participants. Using a two-point prior the probability of molnupiravir being superior to placebo (HR>1) was 75·4%, which was just below our defined threshold of 80% for establishing superiority. Using an uninformative continuous prior, the probability of HR>1 was 94·7%. As an exploratory analysis, the change in viral titre on day 5 (end of treatment) was significantly greater with molnupiravir compared with placebo. A total of 4 participants reported severe adverse events (grade 3+), 3 of whom were in the placebo arm.

**Interpretation:** We found molnupiravir to be well-tolerated, with evidence for high probability of antiviral efficacy in a population of vaccinated and unvaccinated individuals infected with a broad range of viral variants.

**Funding:** Funded by Ridgeback Biotherapeutics and UK National Institute for Health and Care Research infrastructure funding. The AGILE platform infrastructure is supported by the Medical Research Council (grant number MR/V028391/1) and the Wellcome Trust (grant number 221590/Z/20/Z).

## INTRODUCTION

Molnupiravir (EIDD-2801/MK-4482) is the first orally-available directly-acting antiviral licensed for treatment of high-risk individuals with mild-to moderate COVID-19. We have previously reported an optimal dose of 800mg every 12 hours for 5 days of molnupiravir in adults with documented SARS-CoV-2 infection^1^ within AGILE, the UK early-phase platform for experimental COVID-19 therapies [www.agiletrial.net]. MOVe-OUT, a double-blind, placebo-controlled trial in 1433 unvaccinated adults with at least one risk factor for severe COVID-19 illness, reported that molnupiravir decreased clinical progression as judged by hospitalisations and death^2^ - the risk of hospitalization or death at day 29 was 6·8 percentage points lower with molnupiravir than with placebo at the interim analysis and 3·0 percentage points lower in the all-randomized analysis. Preliminary data presented at the 2022 Conference on Retroviruses and Opportunistic Infections from an open-label study from India of 1218 adults also reported a significant reduction in hospitalisations ^3^ (1·5% versus 4·3%) using a generic formulation of molnupiravir. Molnupiravir was also associated with a significantly higher rate of SARS-CoV-2 PCR-negativity after 5 days of treatment (77·1% versus 29·3%), and at days 10 (91·3% versus 70·2%) and 14 (93·9% versus 89·0%). Details on vaccination status of participants were not provided. While these studies have shown molnupiravir to be generally well tolerated, longer term safety continues to be monitored during ongoing clinical studies and pharmacovigilance programs. Although NHC was positive in the Ames test, extensive study of molnupiravir in *in vivo* whole animal mutagenicity assays was reassuring.^4,5^

The shifting epidemiology of SARS-CoV-2 variants globally with the growing prevalence of the Omicron BA.2 lineage gives cause for concern, particularly with an anticipated loss of clinical effect of many monoclonal antibodies. While directly-acting antivirals are expected to remain effective, confirmation of continued efficacy against emerging variants is required. The AGILE platform undertook a seamless phase 1b/2a evaluation of molnupiravir in the UK using a Bayesian adaptive design.^6^ Here, we present the Phase II results in a detailed assessment of clinical outcomes and serial virological responses across a range of SARS-CoV-2 variants, including participants who were vaccinated and unvaccinated.

## METHODS

### Trial design and Oversight

The AGILE CST2 phase II trial (NCT04746183) was designed as a double-blind, randomized, controlled Bayesian adaptive trial in adult early infection in the community with the primary objective to determine the ability of the recommended phase II dose (800mg orally 12 hourly for 5 days) of molnupiravir to improve viral clearance. This was conducted at five UK National Institute for Health and Care Research (NIHR) Clinical Research Facility (CRF) sites (in Liverpool, Manchester, Lancashire, Southampton and London), coordinated by the NIHR Southampton Clinical Trials Unit and sponsored by the University of Liverpool. Eligible participants were men and women aged ≥18 years with PCR-confirmed SARS-CoV-2 infection who were within five days of symptom onset, free of uncontrolled chronic conditions and ambulant in the community with mild or moderate disease. Women of childbearing potential and men were required to use two effective methods of contraception, one of which should be highly effective, throughout the study and for 50 and 100 days thereafter, respectively. Participants were eligible irrespective of whether they were unvaccinated or had received one or multiple UK approved vaccines. Any of the following criteria excluded participants from the study: pregnant or breastfeeding women, stage 4 and 5 (severe) chronic kidney disease, clinically significant liver dysfunction, SpO2 <95% by oximetry or lung disease requiring supplementary oxygen, ALT and/or AST >5 times upper limit of normal, platelets <50 × 10^9^/L, experiencing any grade 3 or above Common Terminology Criteria for Adverse Events (CTCAE version 5), previously reported hepatitis C infection, known allergy to any study medication, or having received any other experimental agents within 30 days of first dose of study drug (use of other co-medications was allowed). All participants provided written informed consent before enrolment.

A total of 180 patients were randomly allocated in a 1:1 ratio using the method of permuted block stratifying for site to either molnupiravir at 800mg twice daily for 10 doses over 5 days or placebo. The study protocol was reviewed and approved by the UK Medicines and Healthcare product Regulatory Agency (MHRA) (EudraCT 2020–001860-27) and West Midlands Edgbaston Research Ethics Committee (20/WM/0136).

### Molnupiravir and placebo

Molnupiravir was provided by Ridgeback Biotherapeutics as 200mg capsules (with matching placebo) administered at 800mg twice daily (morning and evening with water) for ten doses over five days (N.B. days 1-5 or 6 depending on timing of first dose). Participants were randomised to receive molnupiravir or placebo together with standard of care (symptomatic relief including antipyretics) after at least a 2 h fasting period with a 4 h period of observation after the first dose within the clinic and dispensed study drug for twice daily dosing at home. Participants were also required to fast for 1 hour after administration. Participants returned to the clinic on Days 3, 5 and 8, bringing their study medication with them for drug accountability. Participants received a dose of study drug in the clinic on Day 1and the remaining doses of study drug were sent home with the participant for self-administration. Since no specific clinical drug-drug interaction data were available at the time of this trial, the investigators were permitted to apply discretion regarding the use of concomitant medications, guided by the Liverpool COVID-19 Drug Interactions tool (www.covid19-druginteractions.org).

### Efficacy assessment

The primary endpoint was the time from randomisation to negative PCR with an exploratory virological endpoint of change in viral titre. Serial surveillance swabs sampled first from the oropharynx, then nasopharyngeal space were collected in DNA/RNA Shield (Zymo Research #R1100) at screening and again (if the visits were separate) at baseline (day 1), then days 3, 5, 8, 11, 15, 22 & 29. Viral RNA was extracted from samples using Maxwell RSC viral Total Nucleic Acid Purification Kit (# AS1330) according to the manufacturer’s instructions. PCR was carried out (blinded to treatment allocation) using TaqPath COVID-19 RT-PCR Kit (ThermoFisher Scientific, Waltham, Massachusetts), with readings comprising three amplicons: S-gene, N-gene and OFR1 (thresholds were adjusted for each amplicon on each analysis, to give a threshold cycle of 32 with a control of 25 templates per reaction). Time of negativity within an amplicon was determined by the time of the first of two consecutive readings below the limit of detection (cycle threshold of 32 or more) where at least two amplicons were concordant. If all three amplicons differed, the median time to negative PCR was utilised. If only two amplicons were evaluable (e.g. if the third was censored), the later time of the two was utilised. Where only one amplicon was evaluable, time to negative PCR was censored at the last PCR measurement. In the event of S-gene amplification failure, the S-gene was considered censored at day 29 and the rules above applied.

Viral titre was quantified from these swabs as follows: Since approved quantitative standards were not yet commercially available, we developed in-house quantitation based on estimating a viral ‘pseudoconcentration’ (expressed as copies of template per reaction). Swabs dipped into a culture containing 1×10^7^ plaque forming units of SARS-CoV-2 (give strain) were serially diluted to produce a calibration curve. A control known to contain 25 copies per reaction was used to adjust the thresholds on all three templates (S, N and ORF1) to yield a cycle threshold (CT) of 32. Logistic regression was then carried out on each calibration curve giving three different coefficients (these were checked periodically for consistency) which were used to estimate a fold change (from the 25 copy estimate) for any threshold cycle. The average of the estimated titre across the three genes (S, N and ORF1 where available) was calculated and then transformed into log10 values. The change in SARS-CoV-2 viral load in nasopharyngeal swabs was measured by subtracting the log10 estimated titre from baseline. Our primary evaluation was a comparison between arms of the mean reduction (from baseline) in viral load at end-of-treatment (viral load day 5), with secondary evaluations for days 3 and 8.

We undertook an evaluation of the pattern of viral elimination (confirmed as the average value of at least two concordant amplicons), with patients categorised into one of four groups: i) *viral clearance* - stable trajectory of viral load decline to below limit of quantitation, ii) *transient increase in viral titre* - following a viral load reduction, a subsequent rise in titre of at least 0·5 log10 copies, to a level which was maintained or increased on the next consecutive sample iii) *indeterminate* - following a viral load reduction, a subsequent rise in titre which was not confirmed in the next consecutive sample, and iv) *non-evaluable*.

For typing of variants, viral RNA from nasopharyngeal swab was reverse transcribed then sequenced using the EasySeq™ RC-PCR SARS-CoV-2 whole genome sequencing kit (NimaGen, Nijmegen, The Netherlands). Sequence reads were cleaned, trimmed and mapped to the SARS-CoV-2 Wuhan-Hu-1 reference genome (NC_045512.2).^7^ For each sample, genomic variants were called and filtered by quality, with high quality variant calls being used to generate the consensus genome sequence for each sample. The consensus genome sequence was then processed using Pangolin,^8^ a widely used computational tool that assigns the most likely lineage to a given SARS-CoV-2 genome sequence according to the Pango dynamic lineage nomenclature scheme.

Secondary clinical efficacy endpoints were: i) the eleven point WHO Clinical Progression Scale for COVID-19,^9^ ii) the thirty two item NEWS2 score (UK Royal College of Physicians, 2017) measuring acute illness, iii) the FLU-PRO (version 1.2; Leidos Biomedical Research, Maryland, USA) patient self-report of presence and severity of influenza-like symptoms across 6 domains of nose, throat, eyes, chest/respiratory, gastrointestinal and body/system at day 15 and 29. FluPro was recorded at baseline, and on days 15 and 29. Overall survival (time-to-event) was calculated from randomisation to death (any cause), with those still alive censored at last time known to be alive.

### Safety assessments

Safety (and other endpoints) was evaluated at specific time points throughout the trial (Days 1,3,5,8,11,15,22,29 and daily if in hospital) using CTCAE v5 with real time serious adverse event reporting. Data on dose limiting toxicities (DLT – defined as any adverse event of Grade 3 or above using CTCAE version 5 over the first 7 days) were recorded to support the findings of the previous phase I. Time to hospitalisation, hospitalisation rates at day 15 and 29 and duration of any oxygen use/mechanical ventilation were recorded.

### Statistical analysis

The sample size for phase II was based on a time to PCR negativity (censored at 29 days) comparing molnupiravir to placebo. One formal interim analysis (with the independent Data Monitoring and Ethics Committee) was scheduled after 60 participants were enrolled to evaluate futility or efficacy; another interim analysis was added after 120 participants. We used a Bayesian adaptive approach to accelerate decision making, based on a hazard ratio (HR) of achieving PCR negativity with drug compared with placebo. Our primary model utilised a two-point prior based on equal prior probabilities (50%) that the HR was 1·0 (ie no effect) or 1·5 (the threshold of effect judged to be clinically important). We defined *a priori* that if the probability of an HR>1 was in excess of 80% molnupiravir would be recommended for further testing in a larger definitive study. If the probability was less than 0·3 at interim, the study would stop for futility. The maximum sample size of 180 was selected to ensure that the overall probability of concluding that molnupiravir was better than placebo was 0·1 when the hazard ratio was 1 (one-sided type I error accounting for one formal interim analysis) while the power to recommend molnupiravir for further testing was approximately 0·77 if the hazard ratio was 1·5 (equivalent to decreasing the median time to viral clearance from 14 days to 9·3 days or increasing viral clearance after 28 days from 75% to 87·5% with molnupiravir). In addition to the two-point prior, we also used a continuous (uninformative) prior to estimate the probability that the HR was greater than 1 and construct 95% credible intervals. Full details of this methodology are available.^10^

All analyses were intention-to-treat apart from the safety analysis (which included only participants who received at least one dose of the allocated treatment). There was no imputation of missing data, data transformations or adjustment for multiplicity for any of the analyses and results were presented as 2-sided p-values and 95% CIs unless otherwise stated. The primary and secondary analyses were conducted after all 180 participants had been followed through day 29.

The phase II primary analysis involved the comparison of groups on time to viral clearance using a Bayesian Cox proportional hazards model. Time to event data were presented as Kaplan-Meier (KM) curves with secondary analyses comparing treatment arms using simple unadjusted cox regression models. Descriptive analyses of baseline characteristics and other endpoints are summarised using means, medians (from KM curves for time-to-event data) and proportions with corresponding IQRs or 95% CIs as appropriate. Statistical testing for differences between arms utilised two non-parametric evaluations: Initially logrank testing was specified but review by the independent statistical expert in our Data Monitoring and Ethics Committee on 8^th^ November 2021 recommended including the Breslow-Gehan Wilcoxon test as a more sensitive discriminator of differences at early timepoints, anticipated with antiviral therapy. Exploratory subgroup analyses were undertaken based on SARS-CoV-2 variants and whether the participants were vaccinated or unvaccinated.

All analyses were reported according to CONSORT 2010 and the ICH E9 guidelines on Statistical Principles in Clinical Trials. All analyses were carried out in SAS v9.4 and Stata v16 except the Bayesian analyses, which were performed using packages available in R v4.0.2.

## RESULTS

### Trial population

A total of 180 participants underwent randomisation between Sep 8, 2020 and March 16, 2022, 90 randomised to molnupiravir and 90 placebo all of whom were included in the analysis (FIGURE 1).

**FIGURE 1:**
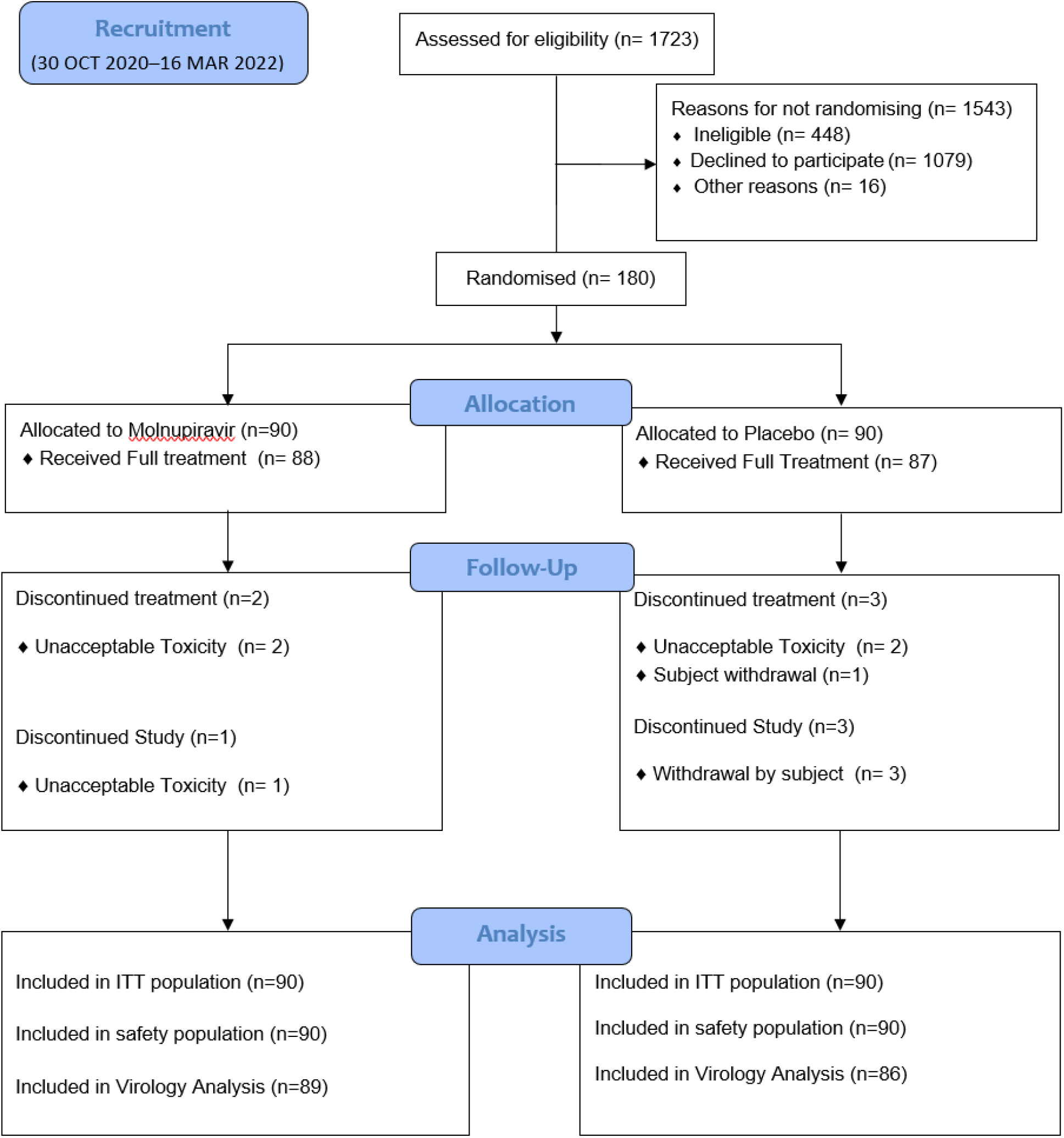
CONSORT.

The baseline characteristics of participants were similar across the molnupiravir and placebo groups with an overall median age of 43, 57% (103/180) female, 100% (180/180) having a WHO COVID progression score of 2 (ambulatory mild disease/symptomatic) and 50% (90/180) having received at least one dose of COVID-19 vaccination at least 14 days before entry into the trial (TABLE 1). The median number of days (range) from symptom onset to randomisation and treatment by the 180 participants was 3 (IQR 3-4) with Delta (40%), Alpha (21%), Omicron (21%), EU1 (16%) and XE (1%) variants represented.

**TABLE 1:**
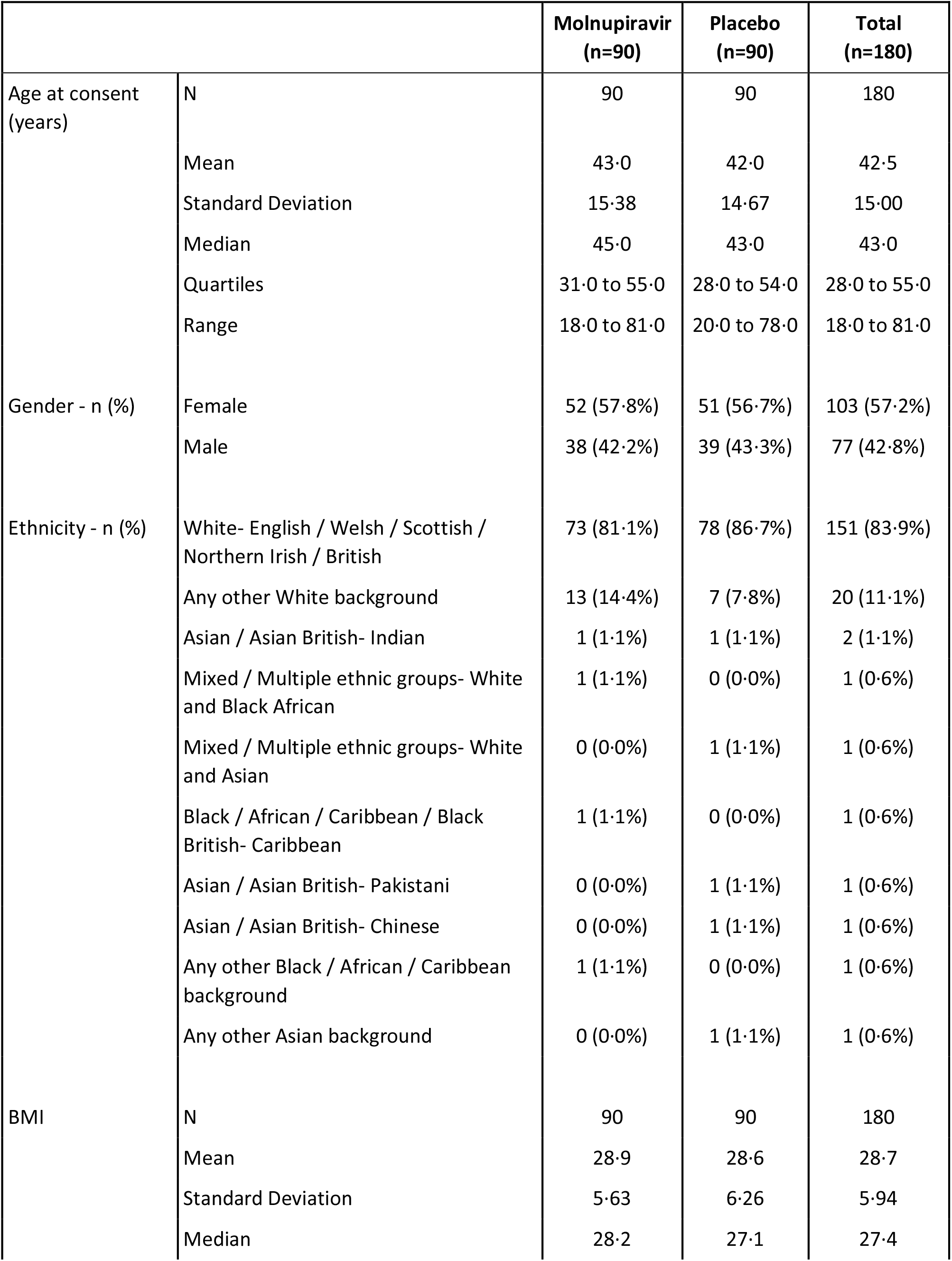
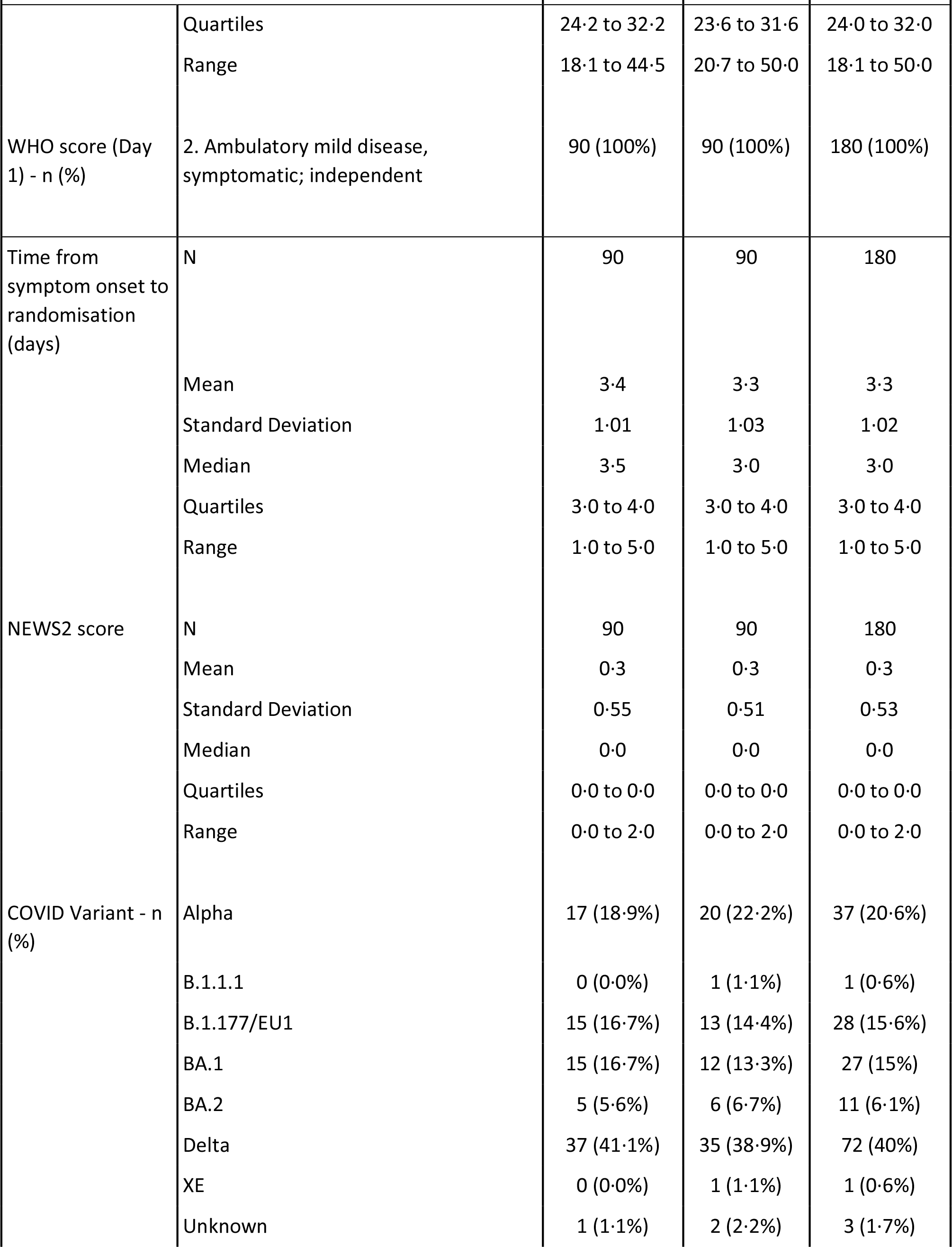
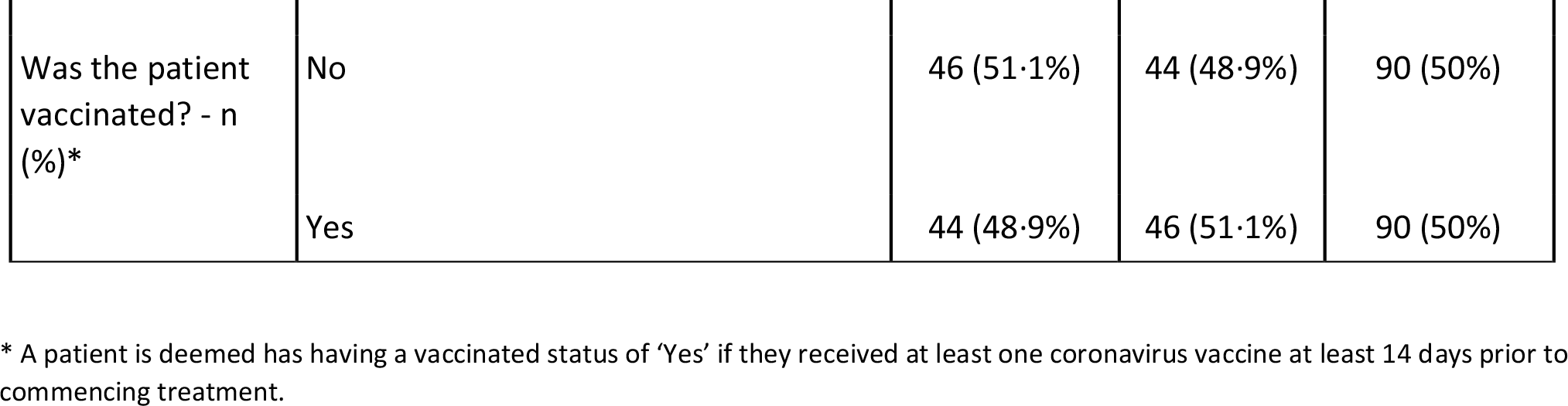
Participant characteristics at baseline (ITT population)

All 180 participants received at least one dose of treatment with 97·8% (88/90) and 96·7% (87/90) completing the full treatment of molnupiravir and placebo respectively. All participants received their first dose, with both arms receiving a median of 10 doses (IQR 10-10) over a median 5 days on treatment (IQR 5-6). A total of 5/180 participants ended treatment early - 2 in each the molnupiravir and placebo arms due to adverse events and 1 in the placebo arm due to participant withdrawal.

### Virological efficacy outcomes

For the primary analysis, molnupiravir was associated with faster median time to negative PCR of 8 days (95% CI 8-9) versus 11 days (95% CI 10-11) for placebo (log-rank p-value 0·07, Breslow-Gehan p-value 0·03 - see TABLE 2 and FIGURE 2). The Bayesian Cox model based on a two-point prior gave a probability of the HR > 1 of 75·4% (this was 75.9% at interim analysis for first 60 participants and 53.5% for the first 120 participants) - this just failed to attain the *a priori* threshold of 80% calibrated as the threshold for recommending a candidate for large scale evaluation. However, the sensitivity analysis using non-informative continuous priors gave a corresponding probability of the HR being greater than 1 of 94·7% (FIGURE S1), with an estimated HR of 1·30; 95% credible interval-CrI (0·92-1·71). The exploratory analysis of time to PCR negativity by vaccination status, variant, gender and ethnicity are shown in FIGURE 3. Numbers are too small for statistical evaluation; efficacy appeared similar in vaccinated participants relative to unvaccinated with nominally greater effect in those who were vaccinated.

**Table 2:** Time to negative PCR - primary analysis

| All patients | Molnupiravir (n=90) | Placebo (n=90) |
| --- | --- | --- |
| **Median time (days) to negative PCR (95% CI)** | 8 (8, 9) | 11 (10, 11) |
| **Negative PCR (n (%)) / (95% CI) at:** |  |  |
| **Day 15** |  |  |
| Number of patients | 77 (85.6%) | 73 (81.1%) |
| Estimate from KM plot | 86.5% (78.6%, 92.6%) | 84.2% (75.6%, 91.0%) |
| **Day 29** |  |  |
| Number of patients | 85 (94.4%) | 79 (87.8%) |
| Estimate from KM plot | 95.5% (89.7%, 98.5%) | 91.5% (84.3%, 96.2%) |
| **Median time (days) to negative PCR (95% CI)** |  |  |
|  | (n=17) | (n=20) |
| Alpha | 11 (5, 22) | 11 (11, 15) |
|  | (n=15) | (n=13) |
| B.1.177/EU1 | 8 (5, 8) | 11 (8, 15) |
|  | (n=15) | (n=12) |
| BA.1 | 8 (5, 8) | 11 (5, 14) |
|  | (n=5) | (n=6) |
| BA.2 | 8 (3, n/a) | 15 (7, n/a) |
|  | (n=37) | (n=35) |
| Delta | 11 (8, 12) | 10 (8, 11) |

**FIGURE 2:**
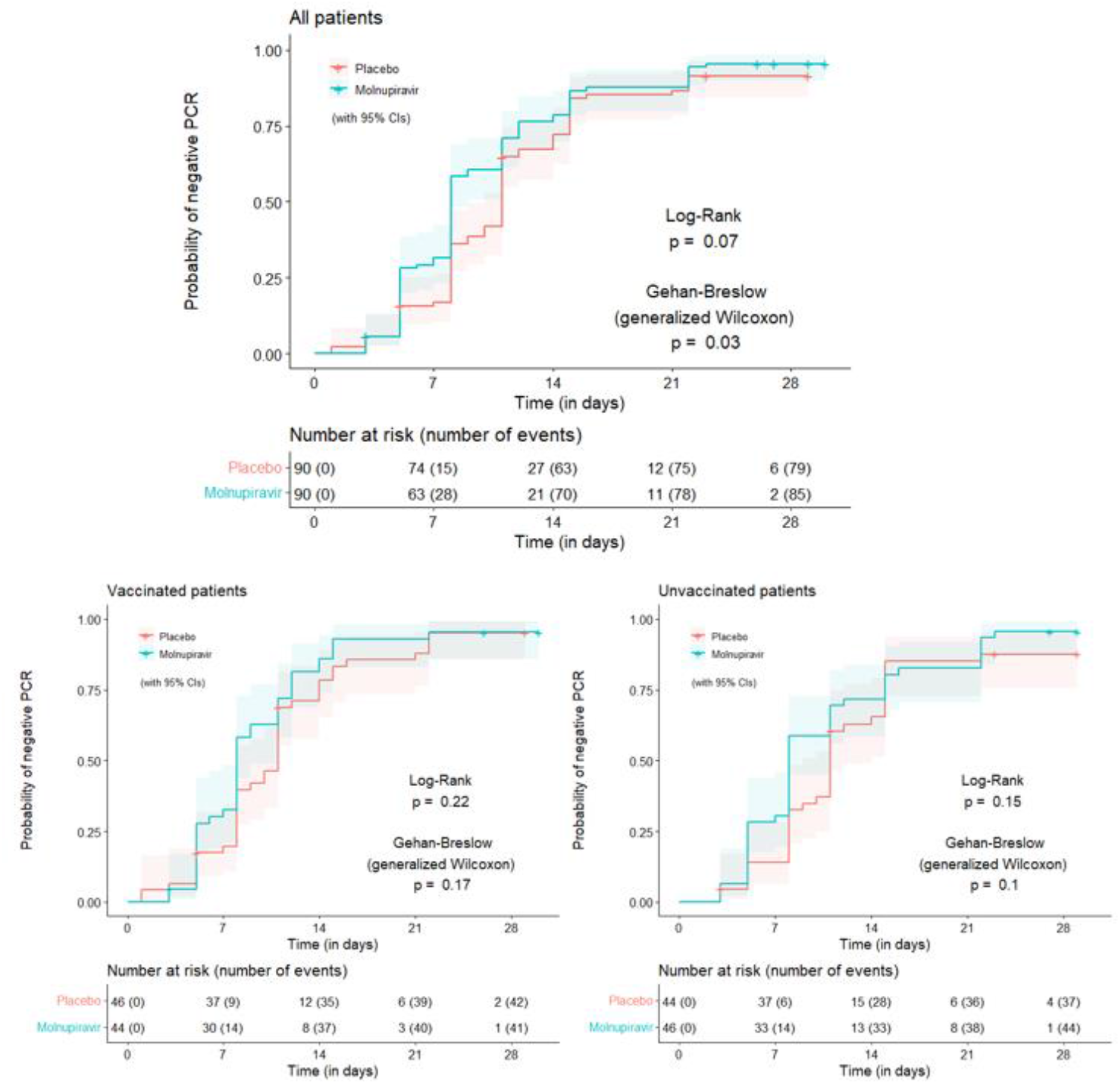
Time to negative PCR (All participants, Vaccinated and Unvaccinated participants)

**FIGURE 3:**
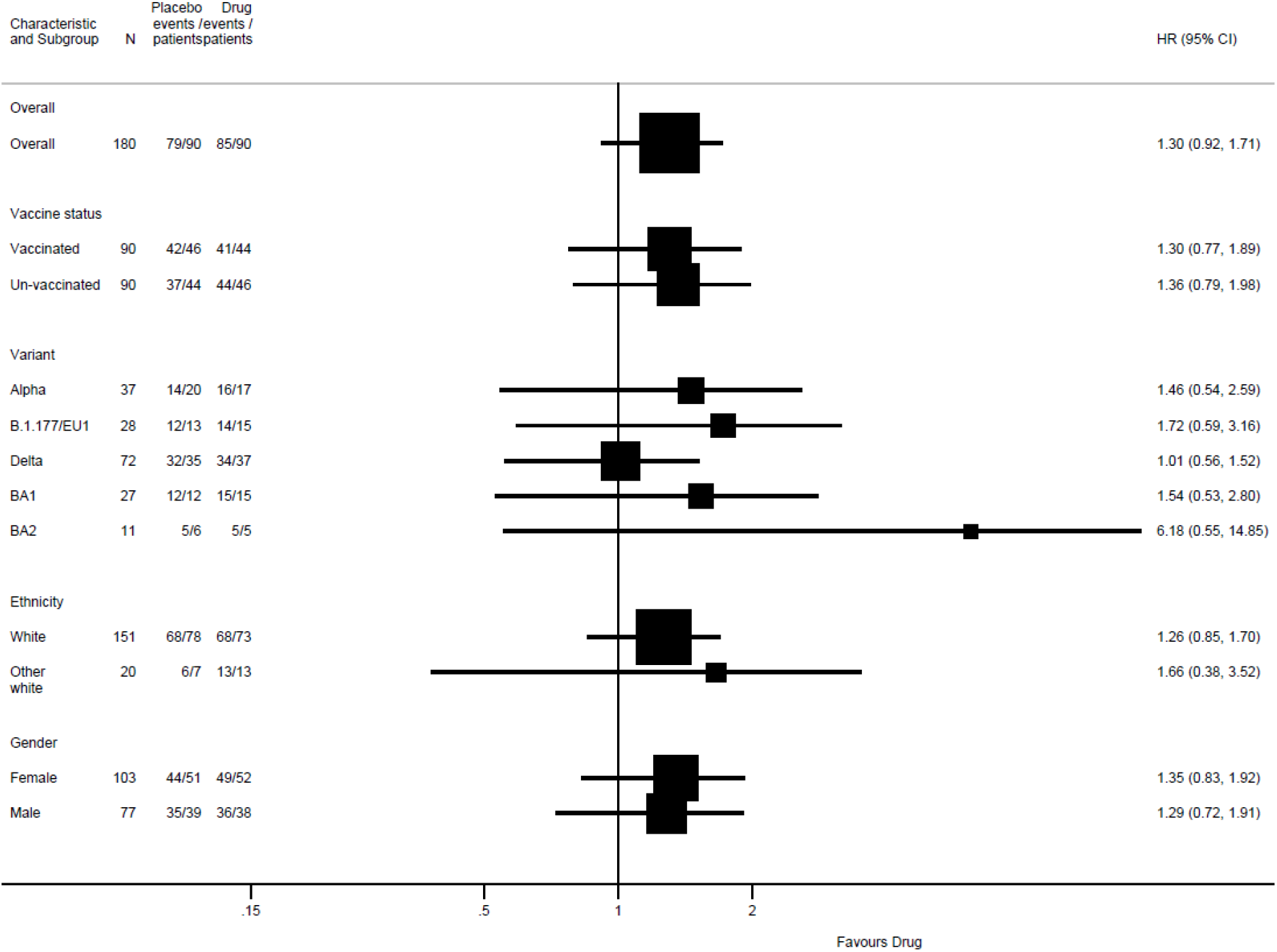
Forest plot **f**or time to negative PCR.

We evaluated change in viral titre as an exploratory efficacy endpoint. Mean (sd) baseline titres for molnupiravir and placebo arms were 7·1 (2·69) and 7·4 (3·00) log10 copies respectively. At Day 5 of treatment, participants in the molnupiravir arm exhibited a mean reduction in viral load of 4·8 log10 copies, compared with 3·9 log 10 copies (P=0·04). Among vaccinated individuals the reduction in viral titre was 5·4 (molnupiravir arm) versus 4·1 (placebo) log10 copies (p=0·03) and among unvaccinated participants the reduction was 4·2 (molnupiravir) versus 3·6 (placebo) log10 copies (p=0·38).

Different patterns of viral clearance were seen over 29 days. We observed a transient increase in viral titre in 7/180 (3·9%; 3 on molnupiravir, 4 placebo). Of the remainder, 74·4% showed a pattern of viral clearance, 18·9% were indeterminate and 2·8% were non-evaluable.

### Clinical efficacy endpoints

No participants died (due to any cause) during the trial. No participants experienced an incidence of SpO2 <92%. No participant in the molnupiravir, compared with four in the placebo arm were hospitalised, one of whom received one day of oxygen. No patient required mechanical ventilation. The WHO Clinical Progression Scale score, NEWS2 assessment score and FluPRO overall scores were similar in each arm at day 15 and 29 (TABLE S1) with 42·4% (73/172) of participants with a WHO score of 0 or 1 by day 15 and 70·5% (122/173) by day 29. Both the NEWS2 (mean 0·3) and FluPRO (mean 0·1 and 0·2) scores were very low at Days 15 and 29.

### Safety outcomes

Of the 180 participants who had received at least one dose and included in the safety analysis 81·1% (73/90) and 75·6% (68/90) of participants experienced an adverse event (i.e. grade 1 and above) in the molnupiravir and placebo arms respectively to Day 29 (TABLE 3). There were a total of 1 and 3 DLTs in the molnupiravir and placebo arms respectively. This comprised 1 participant in the molnupiravir arm (1 grade 3 hypertension) and 3 participants in the placebo arm (8 events including 1 grade 3 vomiting, 1 grade 3 nausea, 1 grade 3 gallstone pancreatitis, 1 grade 3 blood bilirubin increase, 1 grade 3 alanine aminotransferase increase, 1 grade 3 hypocalcemia, 1 grade 3 GGT increase and 1 grade 4 hypomagnesaemia) who experienced a grade 3 or above severity. Two of these four participants were vaccinated and two unvaccinated. Four participants (4.4%) in the placebo arm reported serious adverse events (gallstone pancreatitis, vomiting, hypocalcaemia and hypomagnesemia, and breathlessness) which led to hospitalisation, compared to none in the molnupiravir arm.

**Table 3:**
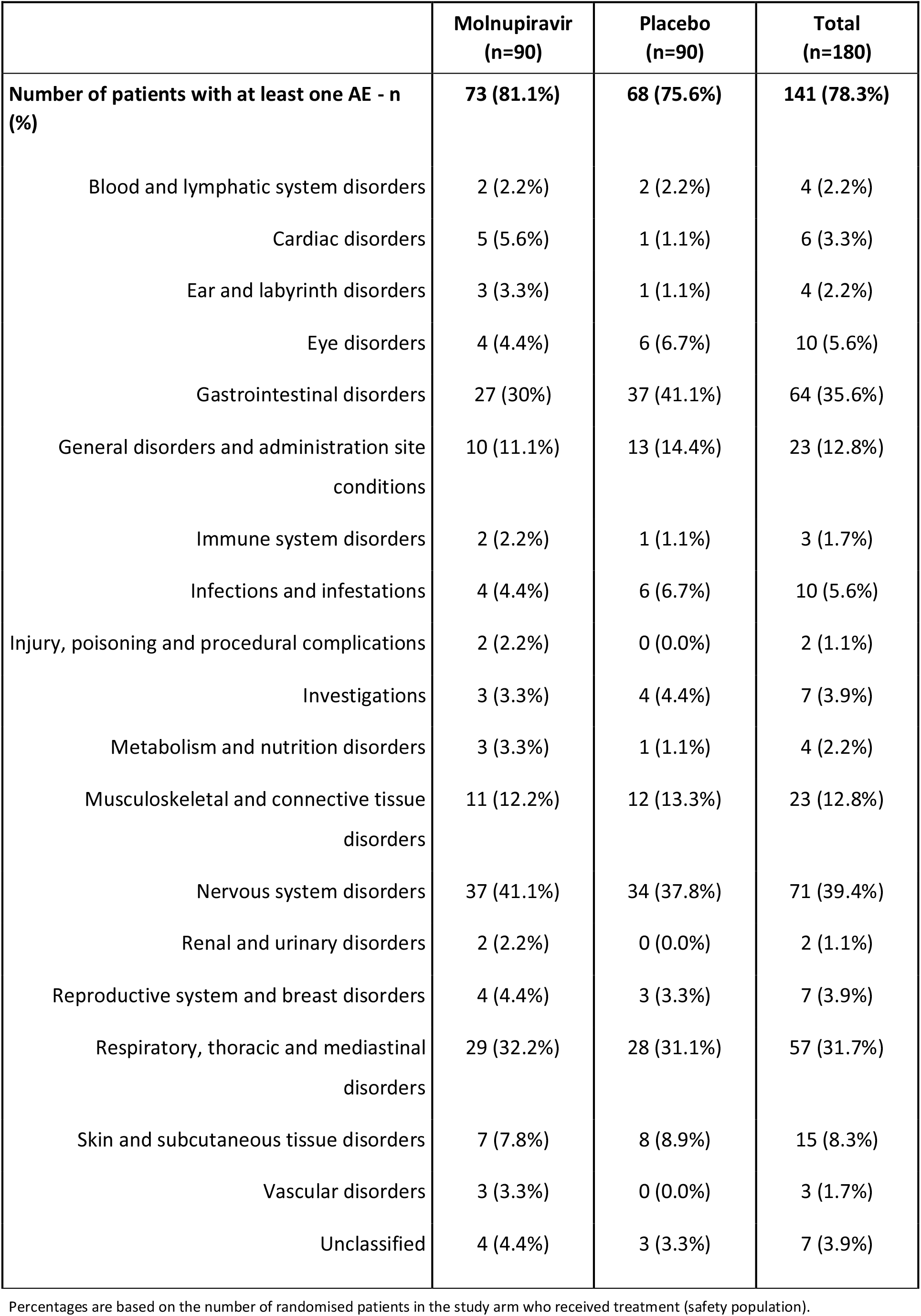
Overall Adverse Events by System Organ Class.

## DISCUSSION

Molnupiravir received conditional marketing authorisation from the UK Medicines and Healthcare Products Regulatory Agency, and early use authorisation from the US Food and Drug Administration based on data from the MOVe-OUT study in unvaccinated individuals at high risk of severe disease, who were infected with variants in circulation between May-October 2021.^2^ MOVe-OUT observed good tolerability of molnupiravir, and reported an approximately 50% reduction in hospitalisations and death at interim evaluation, falling to around 30% after all 1433 patients were analysed. Evaluation of virological response by SARS-CoV-2 variant, and in vaccinated patients within a randomised controlled trial has not been previously published.

In our phase II study, we observed faster time to PCR-negativity with molnupiravir compared to placebo (8 versus 11 days; p=0·07 by logrank test, and p=0·03 using the Breslow-Gehan test which was used to better discriminate early changes between groups). AGILE employs a Bayesian framework to facilitate decision-making. Using a two-point prior approach, the probability of a HR>1 for PCR-negativity compared to placebo was 75·4% (just failing to achieve the 80% target we had set *a priori* as a threshold for a clear decision to progress clinical evaluation). However, when a continuous, non-informative prior was used, molnupiravir was predicted to have a high (94·7%) likelihood of a HR>1 for PCR-negativity compared with placebo. Subgroup analyses lacked sufficient power for statistical comparison. There was no obvious loss of effect by vaccination status (50% vaccinated) or with omicron variant (21%; compared with other variants or overall response).

Participants receiving molnupiravir also had a significantly greater relative reduction in viral load at the end of treatment (five days) compared with placebo (−0·9 log pseudocopies; p=0·04). This significant difference was retained when evaluating only vaccinated participants, but was not maintained when evaluating only unvaccinated patients. The transient rise in viral titres observed in a minority of patients followed out to day 29 was not associated with any return or worsening of clinical symptoms, and likely reflects the natural history of viral infection.

Our study has several limitations and strengths. The number of participants in this phase II trial was limited, and lacked power to detect differences in clinical events such as hospitalisation and death. The clinical tools utilised (W HO clinical progression score, and FLUPRO instruments) lacked sensitivity to detect small changes in an ambulatory cohort of patients. Caution should be exercised when interpreting outcomes for subgroups where sample size is small. We did not culture virus, and time to PCR-negativity may be an insensitive marker for tracking any effect of molnupiravir, given its known mechanism of action. We were however able to undertake rich serial sampling of nasopharyngeal swabs to characterise viral elimination rates and trajectories. Our study also included data in vaccinated participants, and with the Omicron variant, which have been key gaps in clinical evidence for molnupiravir thus far. The efficacy of molnupiravir against these newer variants will be evaluated in the PANORAMIC trial^11^ which is the largest randomised evaluation of molnupiravir to date.

We found molnupiravir to be well-tolerated over 29 days of assessment. Most adverse events were mild, and likely related to COVID-19 with a low frequency of severe adverse events (1·1% grade 3 or higher and no serious adverse events observed in the molnupiravir arm). No hospitalisations occurred in the molnupiravir arm, compared with 4 hospitalisations in the placebo arm (1 case each of severe vomiting, gallstone pancreatitis, severe electrolyte disturbance with hypomagnesemia and hypocalcaemia, and dyspnoea requiring oxygen therapy).

In summary, our study is consistent with previous findings of antiviral efficacy of molnupiravir. Although numbers were small, this efficacy was also retained in vaccinated individuals. A broad range of infections with SARS-CoV-2 variants were included, including participants with Omicron. These data from our randomised trial are consistent with preliminary observational data from Hong Kong^12^ which collectively add to growing evidence of generalisability of trial findings to newer variants.

## Research in Context

### Evidence before the study

Molnupiravir was the first orally-administered directly-acting antiviral for treatment of SARS-CoV-2 infection, which gained conditional marketing authorisation from the UK Medicines and Healthcare Regulatory Agency in November 2021, and early use authorisation from the US Food and Drugs Administration in December 2021. These approvals were based on interim analysis of the MOVe-Out study where 775 unvaccinated adults at high risk of developing severe COVID-19 disease were randomised to receive 5 days of molnupiravir or placebo; molnupiravir was associated with a significant reduction in hospitalisations and deaths. We searched PubMed from the inception of the database to 30^th^ June 2022, using the search terms “SARS-CoV-2”, “randomised trial” and “molnupiravir”. A phase 2a trial reported faster viral clearance with molnupiravir compared to placebo; however only 53 participants received the currently approved dose of molnupiravir. The full dataset from MOVe-Out (including all 1433 participants) showed an absolute difference in hospitalisations and deaths of 3% (compared with 6.8% at interim analysis). Both these studies did not include vaccinated participants, and were undertaken before the Omicron variants predominated. Preliminary data from India observed a lower incidence of hospitalisations in 1218 adults with mild COVID-19 who randomised to receive open label generic molnupiravir plus standard of care (which in addition to antipyretics included ivermectin and budesonide) versus the latter alone. No details on variants or vaccination status were provided. There is a need to confirm these previous findings in vaccinated individuals infected with contemporary SARS-CoV-2 variants.

### Added value of this study

Our data are derived from rich serial sampling within a stringent randomised, placebo-controlled trial which has enabled differences in time to PCR-negativity and changes in viral titre to be evaluated. The evidence for high probability of antiviral efficacy of molnupiravir in a population of vaccinated and unvaccinated individuals infected with a broad range of viral variants is in keeping with previous observations of viral clearance.

### Implications of all the available evidence

We have shown here continued evidence for the antiviral effect of molnupiravir. Definitive evidence for its clinical effectiveness in a highly vaccinated population is anticipated from the UK’s PANORAMIC trial, which has included over 25,000 participants and which is due to report later in 2022.

## Data Availability

All data produced in the present study are available upon reasonable request to the authors

## AGILE CST-2 Study Group

<u>AGILE Trial Steering Committee</u>

Nicholas Paton (chair), Fred Hayden, Janet Darbyshire, Amy Lucas, Ulrika Lorch

<u>AGILE independent Data Monitoring and Ethics Committee</u>

Andrew Freedman (chair), Richard Knight, Stevan Julious

<u>Liverpool School of Tropical Medicine</u>

Rachel Byrne, Ana Cubas Atienzar, Jayne Jones, Chris Williams

<u>Southampton Clinical Trials Unit</u>

Anna Song, Jan Dixon, Anja Alexandersson, Parys Hatchard, Josh Northey, Emma Tilt

<u>University of Lancaster</u>

Andrew Titman

<u>University of Liverpool</u>

Ale Doce Carracedo, Vatsi Chandran Gorner, Andrea Davies, Louis Woodhouse, Nicola Carlucci

<u>NIHR Liverpool Clinical Research Facility</u>

Emmanuel Okenyi, Marcin Bula, Kate Dodd, Jennifer Gibney, Lesley Dry, Zalina Rashid Gardner, Amin Sammour, Christine Cole, Tim Rowland, Maria Tsakiroglu, Vincent Yip, Rostam Osanlou, Anna Stewart

<u>NIHR Manchester Clinical Research Facility</u>

Ben Parker, Tolga Turgut, Afshan Ahmed, Kay Starkey, Sujamole Subin, Jennifer Stockdale, Lisa Herring

<u>NIHR Southampton Clinical Research Facility</u>

Jonathan Baker, Abigail Oliver, Mihaela Pacurar, Dan Owens, Alistair Munro, Gavin Babbage, Saul Faust, Matthew Harvey, Danny Pratt

<u>NIHR Kings Clinical Research Facility</u>

Deepak Nagra

<u>NIHR Lancashire Clinical Research Facility</u>

Aashish Vyas

## Contributors

SK, GG, TJ, RF GS, SE, PM contributed to study design. SK, GG, TJ, GS, SE, PM contributed to data analysis and interpretation. RF, SA, CJE, DH, JP led clinical conduct as principal investigators of the clinical sites. TF, LW, AB, DC, RL participated in clinical assessment and data collection. WG, LLL, KB, VS, TE, CH, JAH, IDB contributed to study bioanalysis. MT, DGL, MJ, JC, EK, ND, HR CW, CM contributed to study management and execution. WH, WP contributed preclinical and safety data on molnupiravir. The manuscript was written by the authors, with SK and GG as the overall lead authors. No one who is not an author contributed to writing the manuscript. All authors had full access to the data and GS and SE directly accessed and verified the underlying data reported in the manuscript. The authors assume responsibility for the accuracy and completeness of the data and for the fidelity of the trial to the protocol.

## Data Sharing Statement

The AGILE Trial Steering Committee will consider all reasonable requests by health-care providers, investigators, and researchers to provide anonymised data to address specific scientific or clinical objectives. The AGILE investigators are committed to reviewing requests from researchers for access to clinical trial protocols, de-identified patient-level clinical trial data, and study-level clinical trial data. Data will be assigned a DOI through deposition in the University of Liverpool Research Data Catalogue and shared under a Data Transfer agreement (or equivalent e.g. as part of a research collaboration agreement or confidentiality disclosure agreement).

## Acknowledgements

Ridgeback Biotherapeutics provided funding for AGILE CST-2 and supply of molnupiravir and matched placebo. The AGILE platform infrastructure is supported by the Medical Research Council (grant number MR/V028391/1) and the Wellcome Trust (grant number 221590/Z/20/Z). UK National Institute of Healthcare Research core funding supported Southampton Clinical Trials Unit and Clinical Research Facilities in Liverpool, Southampton, Manchester, Lancashire and Kings College Hospital. JAH is funded by the U.S. Food and Drug Administration Medical Countermeasures Initiative contract (75F40120C00085). The article reflects the views of the authors and does not represent the views or policies of the FDA. Additional support from the NIHR Health Protection Research Unit in Emerging and Zoonotic Infections (award 200907) and the UK Medical Research Council (MR/W005611/1). TJ also received funding from UK Medical Research Council (MC_UU_00002/14). The Bayesian adaptive analysis method utilised in this study was co-designed by PM who is supported by the National Institute for Health Research through NIHR Advanced Fellowship (NIHR300576).

We are deeply grateful to all participants for taking part in this trial. We are also grateful to the following:

<u>Southampton Clinical Trials Unit</u>

Sara Yates, Kiera Fines, Megan Lawrence, Andrea Corkhill

<u>University of Lancaster</u>

Helen Berrington

<u>NIHR Lancashire Clinical Research Facility</u>

Jacqueline Bramley, Rosalind Szurko, Mathew Anuj

<u>University of Oxford</u>

Ling-Pei Ho

<u>Southampton City Council</u>

Vicky Toomey

<u>Liverpool City Council</u>

Paula Parvulescu, Sophie Kelly, Callum Rutherford, Richard Jones

